# Seasonal variation in mood and the dynamics of sleep, activity, circadian rhythms, and light

**DOI:** 10.64898/2026.05.03.26352101

**Authors:** Mirim Shin, Emiliana Tonini, Joanne S Carpenter, Mathew Varidel, Alissa Nichles, Natalia Zmicerevska, Elizabeth Phung, Connie Janiszewski, Minji Park, Daniel Froggatt, Dhyana Hanlon, Amira Chami, Min K Chong, Haley LaMonica, Frank Iorfino, Angus C Burns, Sean W Cain, Sun Jung Kang, Vadim Zipunnikov, Wei Guo, Debangan Dey, Andrew Leroux, Kathleen R Merikangas, Elizabeth M Scott, Ian B Hickie, Jacob J Crouse

## Abstract

**Background:** Mood symptoms vary seasonally, yet the underlying mechanisms remain unclear. We tested whether wearable-derived sleep, activity, circadian, and light exposure patterns mediate seasonal effects on mood in youth with emerging mood disorders.

**Methods:** We analysed 733 observation periods from 422 Australian youth (mean age 24.3±5.5 years; 63% female) attending early-intervention mental health services. Each observation comprised a clinical assessment paired with ≥5 valid days of GENEActiv wrist actigraphy. Season was modelled using sine-cosine functions of day-of-year. Sleep, activity, and circadian features were reduced using Joint and Individual Variation Explained, and light exposure features were reduced via principal components analysis. Linear mixed-effects models tested seasonal effects on depressive, psychiatric, manic, and functional outcomes. Mediation was examined using Sobel screening followed by cluster bootstrapping (1,000 iterations).

**Results:** Depressive (β=−0.67, p=0.023) and negative symptoms (β=−0.17, p=0.041) peaked in winter, whereas manic symptoms peaked in autumn (β=0.24, p=0.018). Reduced day-to-day variability in moderate-to-bright ambient light exposure (fewer transitions to brighter environments) mediated winter increases in depressive (indirect β=−0.06, p=0.006) and negative symptoms (indirect β=−0.05, p<0.001). Higher activity levels partially mediated season’s effect on depressive symptoms (indirect β=−0.010, p=0.032). Extended sleep with nocturnal activity mediated season’s effect on negative symptoms (indirect β=−0.02, p=0.001). No mediators emerged for manic symptoms.

**Conclusions:** Light exposure variability—reflecting constrained engagement with brighter environments during winter—emerged as the dominant mediator of seasonal mood worsening in Australian youth, with smaller contributions from sleep-activity-circadian patterns. These findings identify daily light variability as a promising, modifiable target for intervention.

## INTRODUCTION

Mood disorders such as depressive and bipolar disorders are among the most costly and debilitating illnesses in Australia [1] and worldwide [2]. Considerable evidence suggests that mood symptoms exhibit seasonal variation, with seasonal patterns observed across the mood disorder spectrum. Seasonal variation in mood encompasses a broad spectrum of phenomena, from mild within-person fluctuations across the year to the recurrent, impairing episode patterns described as Seasonal Affective Disorder (SAD)[3], although the validity of SAD as a discrete diagnostic entity, as opposed to a transdiagnostic dimensional trait, remains debated [4].

In clinical samples, depressive episodes are more prevalent in winter whereas manic episodes peak in summer [5], with hospitalisations for manic episodes showing spring and summer peaks, and (hypo)manic episodes showing a minor autumn peak [6, 7]. Population-based findings are more complex: a Dutch sample reported lower depressive symptoms and higher positive affect in spring [8], and suicides consistently peak in spring [9, 10]. Higher outdoor spring temperatures has also been associated with improved mood, energy, and sleep quality in bipolar and major depressive disorders [11]. These divergent patterns likely reflect different expressions of seasonal sensitivity across populations and measurement contexts, but the overall pattern of seasonal mood variation—and its potential for guiding the timing and type of self-care and clinical management strategies [12]—has prompted investigation into its underlying mechanisms.

Individual sensitivity to weather and climate changes has been proposed as a potential underlying mechanism [13]. The circadian (body clock) system—which refers to endogenous, self-sustained cycles of ~24 hours that align behaviour and physiological processes with the Earth’s day/night cycle [14]—has been recognised as a particularly compelling pathway driving seasonal mood variation [15]. These biological rhythms adapt to seasonal shifts in day length or photoperiod [16] and influence sleep duration and body temperature [15, 17]. Critically, light exposure is the primary *zeitgeber* (time-giver) for the circadian system, directly entraining the central pacemaker and driving melatonin secretion [18, 19]. Reduced sunlight during colder seasons can lead to decreased serotonin production and altered circadian phase, both of which have been implicated in the worsening of depressive symptoms [20].

Disruptions to circadian rhythms have consistently been associated with both depressive and bipolar disorders [21–23]. “Circadian depression” has been proposed as a potential cross-cutting clinical phenotype associated with circadian disruption, metabolic and inflammatory markers [18, 24, 25]. Despite the established links between season and circadian rhythms, and circadian rhythms and mood, studies that directly test whether circadian rhythm, sleep, and activity patterns mediate seasonal effects on mood symptoms are still lacking.

Most research on seasonality and mood has been conducted in the Northern Hemisphere, with the Southern Hemisphere underrepresented, despite comparable seasonality across hemispheres [26]. Furthermore, the mixed findings may partly reflect reliance on self-reported measures subject to recall bias [27]. Australia, with its four distinct seasons and substantial daylight variation [28], provides an excellent context for investigation; recent Fitbit data confirm that even temperate Australian climates, sleep duration lengthens and sedentary behaviour increases in winter [29].

The use of objective measures, such as wrist-worn accelerometry (actigraphy) to assess sleep-activity-circadian dynamics and light exposure, can reduce subjectivity and strengthen exploration of the mechanisms linking season and mood. Young people with emerging mood disorders are particularly important population for such research. The transition from adolescence to young adulthood is a critical period for the onset of mood disorders [30, 31], and early illness stages could plausibly show different seasonal sensitivity from established chronic illness. Additionally, sleep and circadian disturbances are highly prevalent in youth mental health populations and may represent modifiable treatment targets [23].

This study therefore aimed to: (1) examine seasonal variation in clinical outcomes (depressive, psychiatric, and manic symptoms; social and occupational functioning) in a transdiagnostic sample of Australian youth with emerging mood disorders; and (2) for outcomes showing seasonality, test whether sleep-activity-circadian dynamics and light exposure patterns mediated these effects. We hypothesised that seasonal variation in symptoms and functioning, with mediation by sleep-activity-circadian patterns and/or light exposure patterns.

## METHODS

### Participants and study design

This study pooled data from two transdiagnostic clinical cohort studies of young people accessing mental health services at the University of Sydney’s Brain and Mind Centre (Youth Mental Health Follow-Up Study [YMH]: recruited October 2008-November 2016) and Neurobiology Youth Follow-Up Study [Neurobiology]: recruited April 2021-January 2024). Detailed information about these studies is available in previous publications [32–34]. These studies involved multi-domain assessments across clinical, self-report, neurocognitive, sleep-wake (e.g., actigraphy), and biological (e.g., blood sampling) measures at one or multiple time points. Participants were excluded if they had insufficient English proficiency, an intellectual disability, and in addition, for the YMH study, a history of neurological disease, medical conditions affecting brain function, recent electroconvulsive therapy, or current substance dependence. We note that the Neurobiology study included a pathway to be re-recruited from our prior studies, and as such, the maximum eligible age for the re-recruitment pathway was 40 and for new recruits was 30.

The current analysis included participants with at least 5 valid days of GENEActiv actigraphy data (defined as >16 hours of valid wear time per day, with non-wear identified using GGIR’s non-wear algorithm; valid days were not required to be consecutive) and clinical data from the same visit. Ethics approval was obtained from the University of Sydney Human Research Ethics Committee (2012/1631) for the YMH study and the Sydney Local Health District Human Research Ethics Committee (2020/ETH01272) for the Neurobiology study. All participants provided written informed consent; parental/guardian consent was also obtained for participants under 16 years.

### Measures

#### Clinical assessment

Clinical assessments were administered by trained clinical researchers at the beginning of each actigraphy recording period. We examined Brief Psychiatric Rating Scale (BPRS), a 24-item questionnaire [35, 36], assessing four symptom dimensions (affect, negative, positive, activation); the Young Mania Rating Scale (YMRS) [37], an 11-item clinician-rated measure of manic symptom severity; the Quick Inventory of Depressive Symptomatology (QIDS) [38], assessing the nine DSM-defined symptom domains of a major depressive episode (Neurobiology study only); and the Social and Occupational Functioning Assessment Scale (SOFAS) [39], a clinician-rated 100-point scale of global functioning.

#### Seasonal variables

Season was modelled using sine and cosine transformations of day-of-year to capture continuous periodic seasonal patterns. The start date of each actigraphy recording period was used to index seasonal position for that observation:

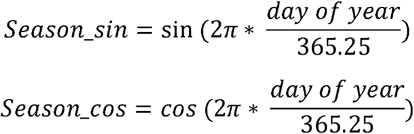

In the Southern Hemisphere, positive cosine values correspond to summer (December-February) and negative values correspond to winter (June-August); the cosine term therefore captures the summer-winter axis. The sine term captures the spring-autumn axis, with positive values corresponding to autumn (March-May) and negative values to spring (September-November). Validation against null and 12-level categorical month models confirmed that the sine-cosine formulation captured seasonal variation parsimoniously (Table S1).

#### Actigraphy data and processing

Detailed actigraphy data processing is presented in the Supplementary Material. Briefly, participants wore GENEActiv accelerometers on the non-dominant wrist for 5-20 consecutive days (median=12 days), with clinical assessments at the start of each actigraphy recording period. Raw accelerometry was processed using GGIR (v3.0.5) [40], followed by mMARCH.AC (v2.9.2) [41]. Light variables were derived from the GGIR Part 5 epoch-level (minute-level) output. Accelerometry and light data were inherently concurrent, as both are recorded simultaneously by the device. To capture both shared and domain-specific information, the Joint and Individual Variation Explained method (JIVE) method [42] was applied to 28 actigraphy-derived variables spanning sleep (7), physical activity (7), and circadian rhythm (14) domains (Table S2). Component loadings and structure are reported in the Results and Table S3.

Light parameters were analysed separately because they represent a distinct source of circadian input. Raw light data are shown descriptively in Figure 1. Following a previous study [43], light exposure was characterised across seven lux thresholds (10, 20, 50, 100, 300, 500, 1,000 lux) using three metrics: *Light Regularity Index* (LRI; day-to-day consistency), *mean light timing* (MLiT) and *photoperiod interval* (Supplementary Materials). Principal Component Analysis (PCA) with oblimin rotation was applied to these 21 variables. After quality control, complete data were available for 491 observation periods from 254 participants (67.0%). Because missingness in light data depends on time- and context-specific environmental factors (e.g., weather conditions, location, and individual behaviour) not captured by study covariates, analyses were restricted to complete cases. Sensitivity analyses confirmed PCA solution robustness to repeated observations and to excluding the 1,000-lux variables (Supplementary Materials).

**Figure 1.**
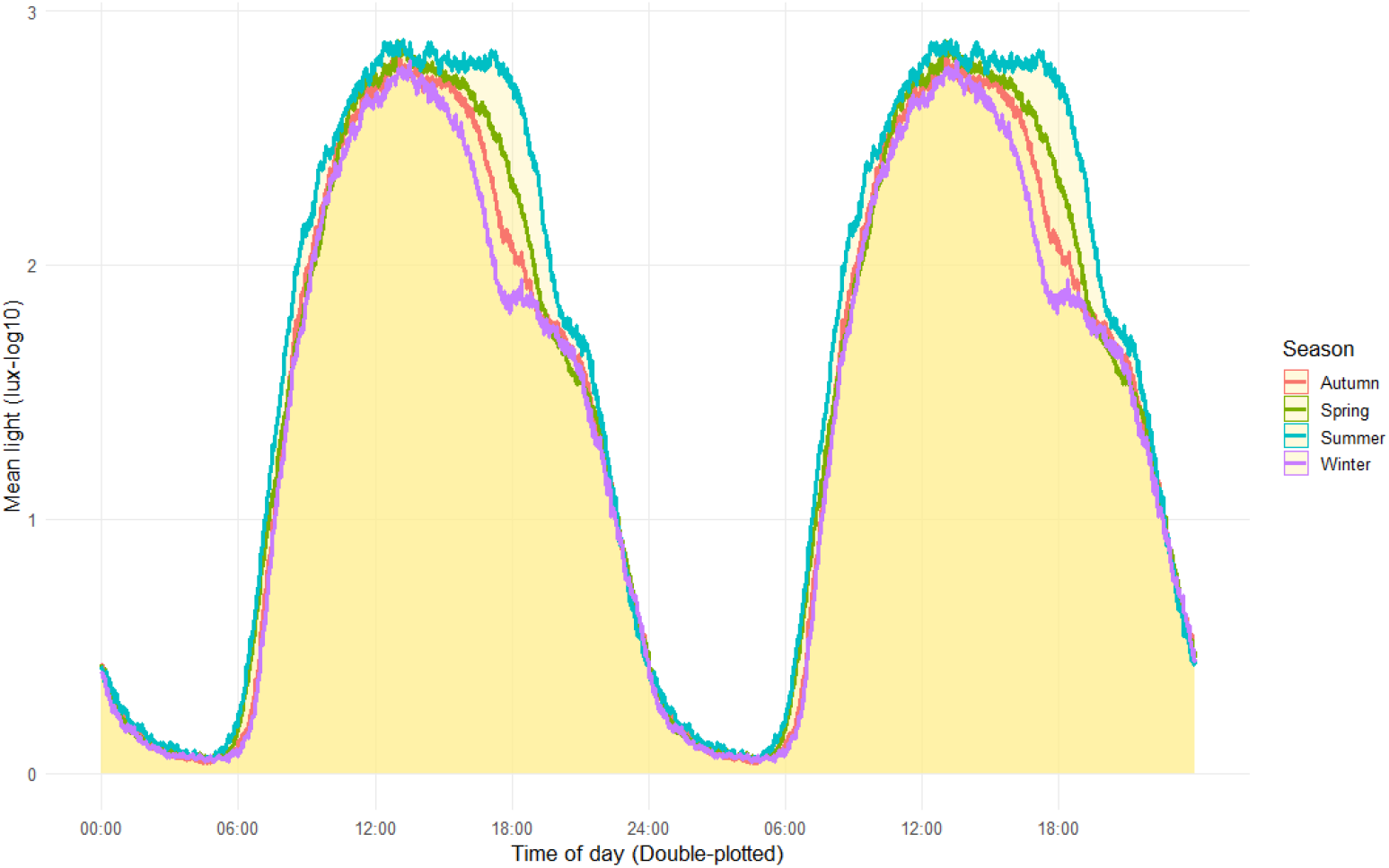
Seasonal variation in ambient light exposure recorded by actigraphy sensors. **Note:** Double-plotted mean daily light exposure [log_10_ (lux+1)] by Southern Hemisphere seasons, derived from GENEActiv wrist-worn sensors across the full study sample. Curves represent group means across 1-minute epochs. The GENEActiv sensor saturates at 3,000 lux, so outdoor daylight is underestimated.

### Statistical Analysis

All analyses were conducted in R v4.5.0 [44] using the lme4 package for linear mixed-effects models [45]. As exploratory analyses, no corrections for multiple comparisons were applied. All models included age and sex as covariates and a participant-level random intercept.

Aim 1: Seasonal effects on seven clinical outcomes (QIDS, YMRS, BPRS subscales [affect, positive, negative, activation]; SOFAS) were tested using linear mixed-effects models with both sine and cosine seasonal terms entered simultaneously. Evidence of seasonality was defined as p<0.05 for either seasonal component. Full model specification is in Supplementary Materials, and estimated seasonal trajectories for significant outcomes are displayed alongside observed monthly means in Figure 2.

**Figure 2.**
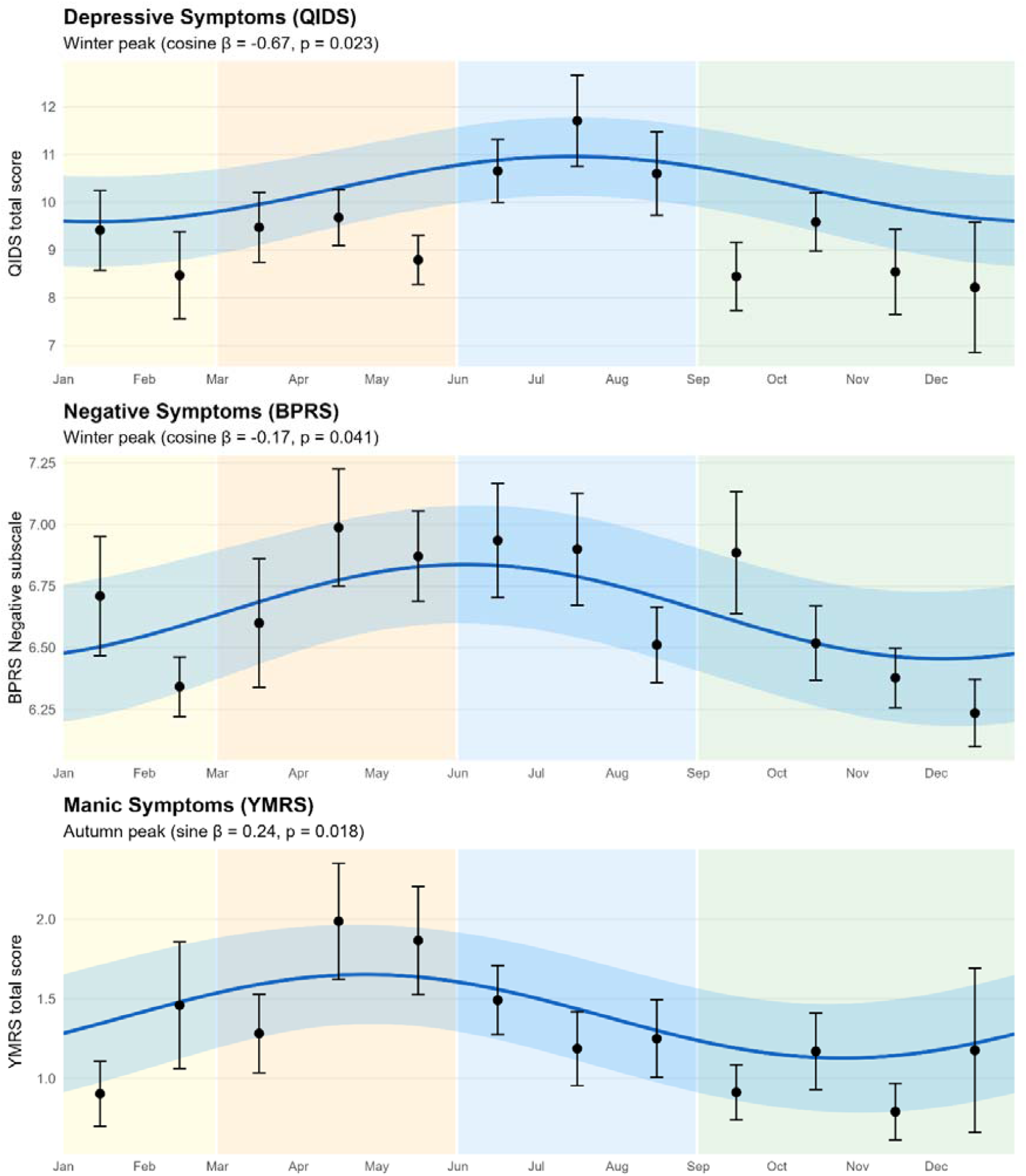
Model-predicted seasonal trajectories for significant clinical outcomes **Note**: Blue line = population-level sine/cosine model prediction; shaded ribbon = 95% confidence interval. Black points = observed monthly means ± SE. The 12-month categorical model did not significantly improve fit over the sine/cosine model for any outcome (all LRT p > 0.40; see Table S1 for formal comparison). Season shading: yellow = Summer (December-February), orange = Autumn (March-May), blue = Winter (June-August), green = Spring (September-November). QIDS = Quick Inventory of Depressive Symptomatology; BPRS = Brief Psychiatric Rating Scale; YMRS = Young Mania Rating Scale.

Aim 2: For outcomes with significant seasonality, mediation by sleep-activity-circadian and light exposure variables was tested. Because clinical assessments occurred at the start of each actigraphy period, findings reflect concurrent seasonal patterning rather than strict temporal precedence. All twelve candidate mediators (8 JIVE, 4 PCA) were tested without pre-selection [46, 47]. Indirect effects were screened with the Sobel test (p<0.10); mediators meeting screening underwent cluster bootstrap resampling (1,000 iterations; participant as resampling unit) to derive 95% confidence intervals (Supplementary Materials).

Missing data arose from three sources: 1) QIDS was collected only in the Neurobiology cohort (28.1% missing); 2) BPRS Negative and YMRS had item-level non-response (17.2% and 20.3%); and 3) light PCA scores were unavailable for 33.0% of observations (primarily reflecting non-attainment of the 1,000-lux threshold). Analyses used listwise deletion. Comparison of observation periods with versus without complete light data showed no meaningful clinical differences (Table S7).

## RESULTS

### Sample characteristics

The analysis included 422 participants from the two longitudinal cohort studies conducted between January 2014 and June 2025, contributing 733 observations periods. Each observation period comprised a clinical assessment and concurrent actigraphy recording of at least 5 valid days. Of the 422 participants, 59% contributed a single observation period and 41% contributed multiple observation periods (up to 5); among those with multiple visits, the median follow-up duration was 364 days [IQR 181-725], with age at last visit of 26.0 years [IQR 22.0-30.0]. Participants’ characteristics at each visit are presented in Table S8.

Baseline demographics and clinical characteristics are presented in Table 1. The mean age was 24.3±5.5 years and 63.3% were female. At baseline, participants showed mild-to-moderate depressive symptoms and functional impairment, with low levels of manic symptoms.

**Table 1.** Participant characteristics at baseline (n=422)

| Characteristic | Value (Mean±SD) |
| --- | --- |
| Age (years) | 24.3±5.5 (range: 13-40) |
| Sex (Female, n (%)) | 267 (63.3%) |
| BPRS Affect | 12.8±4.4 |
| BPRS Positive | 7.0±1.9 |
| BPRS Negative | 6.8±1.7 |
| BPRS Activation | 8.7±2.0 |
| QIDS total score | 10.4±5.1 |
| YMRS total score | 1.5±2.1 |
| SOFAS score | 64.9±14.2 |

**Note:** BPRS = Brief Psychiatric Rating Scale; QIDS = Quick Inventory of Depressive Symptomatology (Neurobiology study only); YMRS = Young Mania Rating Scale; SOFAS = Social and Occupational Functioning Assessment Scale.

### Seasonal variation in clinical outcomes

Three clinical outcomes demonstrated significant seasonal variation (Table 2; Figure 2). Depressive symptoms (QIDS total) and negative symptoms (BPRS Negative) exhibited winter peaks, indicated by significant negative cosine coefficients (β=−0.67, p=0.023 and β=−0.17, p=0.041, respectively). Manic symptoms (YMRS) exhibited a significant autumn peak, reflected by a positive sine coefficient (β=0.24, p=0.018). No significant seasonal effects were observed for BPRS affect, positive, or activation subscales, nor for SOFAS scores (functioning). These non-significant outcomes were not examined in mediation analyses.

**Table 2.** Seasonal effects on clinical outcomes (733 observation periods from 422 participants)

| Outcome | Obs (n) | Sine $\beta$ (SE) | Sine p | Cosine $\beta$ (SE) | Cosine p |
| --- | --- | --- | --- | --- | --- |
| BPRS Depressive | 603 (325) | -0.11 (0.21) | 0.610 | -0.10 (0.24) | 0.665 |
| BPRS Positive | 607 (329) | 0.01 (0.08) | 0.857 | 0.02 (0.09) | 0.815 |
| BPRS Negative | 607 (329) | 0.09 (0.07) | 0.220 | **-0.17 (0.08)** | **0.041** |
| BPRS Manic | 606 (328) | -0.06 (0.09) | 0.507 | 0.04 (0.10) | 0.669 |
| QIDS | 527 (266) | -0.15 (0.25) | 0.549 | **-0.67 (0.29)** | **0.023** |
| YMRS | 584 (314) | **0.24 (0.10)** | **0.018** | -0.11 (0.12) | 0.344 |
| SOFAS | 673 (373) | 0.53 (0.46) | 0.247 | 0.14 (0.54) | 0.793 |

**Note:** Obs = number of observation periods; n = number of participants. Bold indicates a significant seasonal effect ( $p<0.05$ for either sine or cosine term). BPRS = Brief Psychiatric Rating Scale; QIDS = Quick Inventory of Depressive Symptomatology (Neurobiology study only); YMRS = Young Mania Rating Scale; SOFAS = Social and Occupational Functioning Assessment Scale.

### Dimension reduction

The JIVE solution comprised three joint components, two sleep-specific components, one physical activity-specific component, and two circadian rhythm-specific components. Joint components captured 41.4-85.4% of variance across the three domains, indicating substantial overlap (Figure S1). Table S3 shows the loadings on each domain, highlighting features with more than 25% of proportional variation. Joint_1 is related to “activity level” (higher scores indicate greater total physical activity, more time in light and moderate-to-vigorous activity, reduced sedentary time, and more frequent transitions to active states). Joint_2 is related to “timing” (higher scores indicate later sleep onset, wake time, and midpoint, alongside later timing of the most active [M10] and least active [L5] periods). Joint_3 captures a pattern of “extended sleep with nocturnal activity” (higher scores indicate longer sleep duration with lower variability, a phase-delayed activity profile with nocturnal emphasis [higher fPC1, reflecting peak activity near midnight and trough in morning hours], and reduced biphasic daytime activity structure [lower fPC4, reflecting attenuated morning and afternoon activity peaks]).

The light exposure PCA yielded four components explaining 84.6% of variance: PC1 (37.4%) captured timing of light exposure across all lux thresholds (higher score represents later light exposure); PC2 (21.0%) captured duration of light exposure (photoperiod interval); PC3 (15.6%) captured regularity of dim indoor light (LRI at 10-100 lux); and PC4 (10.6%) captured regularity of bright ambient light (LRI at 300-1,000 lux).

### Mediation analysis

Sobel screening identified five candidate mediators (p<0.10): three for depressive symptoms (JIVE_Joint_1 [overall activity level], Light_PC1 [light exposure timing], Light_PC4 [bright ambient light regularity]); two for negative symptoms (JIVE_Joint_3 [extended sleep with nocturnal activity], Light_PC4); and no mediators for manic symptoms (Table S9). JIVE-based mediator analyses included 527-607 observation periods; light-based mediator analyses included 483-486 observation periods, reflecting the light data subsample. Following bootstrap testing, four demonstrated significant indirect effects (Figure 3); full results are in Table S9.

**Figure 3.**
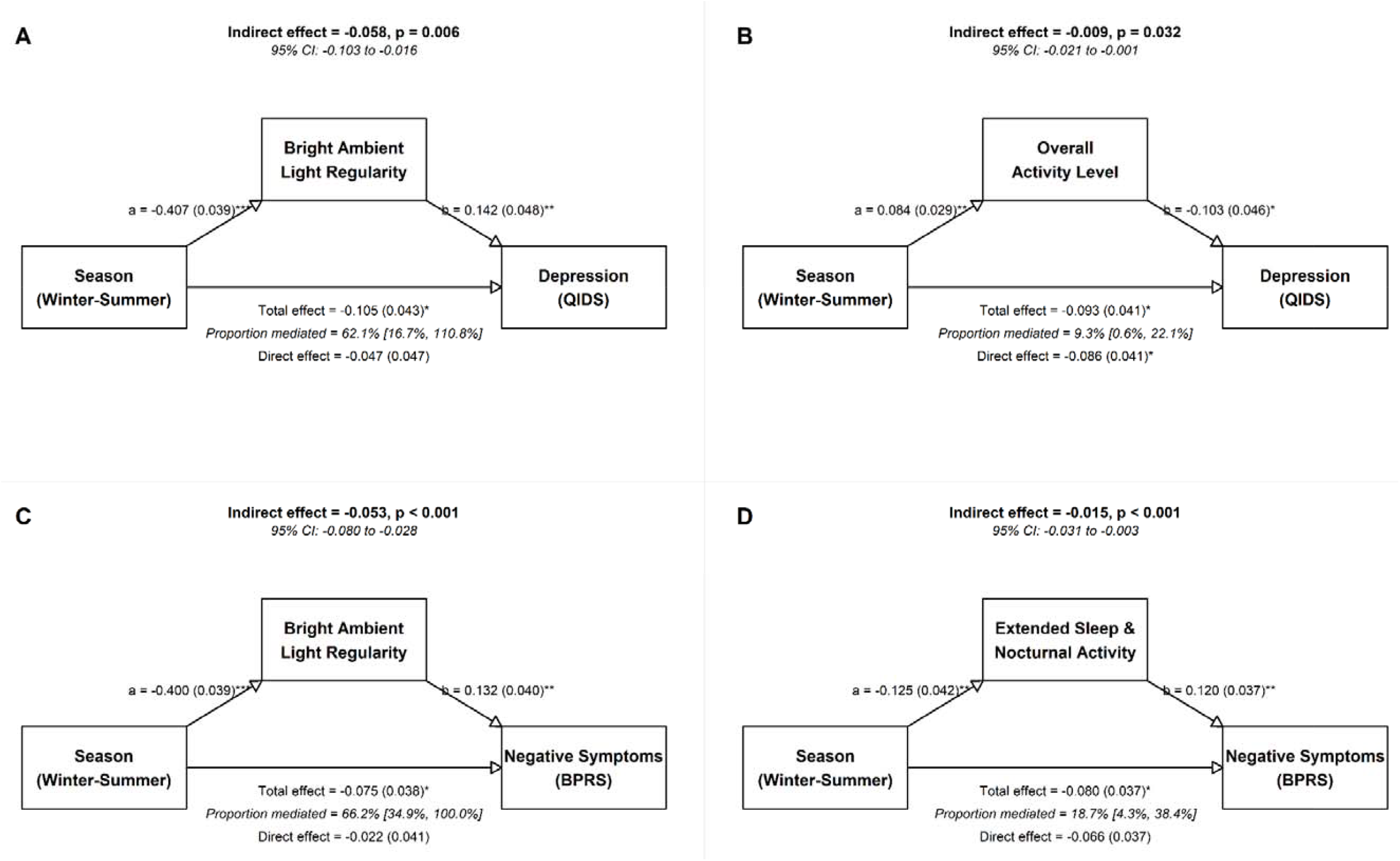
Mediation path diagrams for significant indirect effects of season on mood symptoms (depressive and negative). **Note:** Panels A-B show mediation of seasonal effects on depressive symptoms (QIDS); Panels C-D show mediation of seasonal effects on negative symptoms (BPRS). Season represents the cosine transformation of day-of-year, where negative values correspond to winter and positive values to summer in the Southern Hemisphere. All effects are standardised. Path a=season→mediator; Path b=mediator→outcome. ^#^Bootstrap CIs based on 1,000 iterations with cluster resampling. % Mediated = indirect/total effect; not reported for non-significant mediations. ^*^p<0.05, ^**^p<0.01, ^***^p<0.001.

#### Depressive symptoms

For depressive symptoms (QIDS total), Light_PC4 (regularity at bright ambient light exposure) demonstrated significant mediation of the winter-summer seasonal effect. A lower cosine value (i.e., winter) was associated with increased bright light regularity (path a: β=−0.407, SE=0.039, p<0.001), which in turn was associated with greater depressive symptoms (path b: β=0.142, SE=0.048, p=0.003). The indirect effect was significant (β=−0.058, 95% CI [−0.103, −0.016], p=0.006), accounting for 62.1% of the total seasonal effect. After including Light_PC4 as a mediator, the direct effect of season on depressive symptoms was no longer significant (c’=−0.047, p=0.316), consistent with full mediation. JIVE_Joint_1 (activity level) exhibited partial mediation. Higher cosine value (i.e., summer) was associated with higher activity levels (path a: β=0.084, SE=0.029, p=0.005), which was associated with lower depressive symptoms (path b: β=−0.10, SE=0.046, p=0.026). The indirect effect was significant (β=−0.009, 95% CI [−0.021, −0.001], p=0.032), explaining 9.3% of the total effect. The direct seasonal effect remained significant after accounting for activity (c’=−0.086, p=0.038), indicating partial mediation. Timing of light exposure (Light_PC1) did not significantly mediate the seasonal effect (p=0.072).

#### Negative symptoms

For BPRS negative symptoms, both candidate mediators demonstrated significant mediation. Light_PC4 mediated 66.3% of the total effect. A lower cosine value (i.e., winter) was associated with increased bright light regularity (path a: β=−0.400, SE=0.039, p<0.001), which was associated with greater negative symptoms (path b: β=0.132, SE=0.040, p=0.001). The indirect effect was significant (β=−0.053, 95% CI [−0.080, −0.028], p<0.001), and the direct seasonal effect was reduced to non-significance (c’=−0.022, p=0.587), consistent with full mediation. JIVE_Joint_3 (extended sleep with nocturnal activity) mediated 18.8% of the seasonal effect on negative symptoms. Lower cosine value (i.e., winter) was associated with higher Joint_3 scores reflecting extended sleep with more nocturnal activity (path a: β=−0.125, SE=0.042, p=0.003), which in turn was associated with greater negative symptoms (path b: β=0.120, SE=0.037, p=0.001). The indirect effect was significant (β=−0.015, 95% CI [−0.031, −0.003], p=0.001), and the direct seasonal effect was reduced to non-significance (c’=−0.066, p=0.073).

#### Manic symptoms

No significant mediations were identified for the autumn peak in manic symptoms. None of the sleep-activity-circadian or light exposure variables met criteria for mediation (all Sobel p>0.10).

## DISCUSSION

This study examined seasonal variation in mood symptoms in Australian youth with emerging mood disorders and tested whether objectively measured sleep-activity-circadian patterns and light exposure mediated these effects. As hypothesised, depressive and negative symptoms peaked in winter, whereas manic symptoms peaked in autumn. Other psychiatric symptoms (e.g., positive psychotic symptoms) and social and occupational functioning did not demonstrate seasonal variation. The most salient finding was that day-to-day regularity of moderate-to-bright ambient light exposure accounted for the majority of the winter-related increase in both depressive and negative symptoms. Sleep-activity-circadian patterns showed smaller, outcome-specific mediating effects, and no mediators were identified for manic symptoms.

Day-to-day regularity of moderate-to-bright ambient light (300-1,000 lux; Light_PC4) accounted for the majority of the winter-related increase in both depressive and negative symptoms. Higher regularity—being above or below the same threshold at similar clock times each day [43]—was associated with worse mood, whereas greater irregularity was suggestively protective. This illuminance range sits at the boundary between brighter indoor environments and shaded outdoor light, well below direct outdoor daylight (>10,000 lux). Sensitivity analyses excluding the 1,000-lux threshold yielded a near-identical component dominated by 300-500-lux regularity (Table S6), indicating the finding is not driven by occasional outdoor excursions registering at the highest threshold. Rather than reflecting beneficial regularity of bright outdoor light [48], the component captures a consistent absence of transitions to brighter environments—predominantly staying indoors. We interpret this as evidence that variability in light exposure—what might be characterised as taking “light breaks” from invariant indoor environments—may benefit mood by providing a stronger circadian signal than constant exposure to inadequate indoor light. Consistent with this, Swedish office workers showed approximately eight-fold greater bright daylight exposure in summer than winter [49], and photosensitivity itself appears seasonally modulated, with greater melatonin suppression to moderate light in winter [50], potentially amplifying the impact of prolonged indoor exposure during colder months. Together, seasonal mood vulnerability may stem not only from reduced light availability, but from reduced variability in daily light exposure patterns.

However, device limitations should temper interpretation. The GENEActiv saturates above 3,000 lux, underestimating outdoor daylight (>10,000 lux) [51, 52], and the wrist sensor can be covered by long sleeves in colder months—potentially flattening winter day-to-day variability and biasing seasonal contrasts toward the null. The observed indoor-light association therefore likely represents a conservative estimate. Direct examination of outdoor light was further precluded by 33% missingness at higher thresholds. Sensors capturing the full circadian-relevant dynamic range, worn at less-occluded locations (e.g., chest or eye-level), will be important for future work.

As mediators, sleep-activity-circadian patterns explained smaller proportions of seasonal effects on clinical outcomes. Overall activity level partially mediated only 9.3% of winter-related increases in depressive symptoms, consistent with extensive evidence for reduced physical activity during winter months [53]. More notably, a joint sleep-circadian pattern characterised by extended sleep duration and nocturnal activity mediated nearly one-fifth of the seasonal effect on negative symptoms. This profile resembles hypersomnia and blunted diurnal activation, features commonly observed in circadian-related depressive phenotypes [18, 24, 25], and aligns with evidence that depressive disorders are associated not only with reduced activity but with dampened circadian amplitude and altered activity timing [54, 55]. Importantly, these sleep-activity effects were smaller than those observed for light exposure regularity, suggesting that while sleep and activity rhythms contribute to seasonal symptom expression, they may do so partly downstream of, or in parallel with, light-driven behavioural constraints.

These findings suggest intervention targets. Behavioural and temporal patterns of light exposure, rather than total light alone, may mitigate seasonal mood worsening. Encouraging regular daily departures from uniformly lit indoor environments—through outdoor activity (e.g., walks outdoors) or exposure to substantially brighter environments (e.g., sitting by a window)—may constitute beneficial “light breaks”. Promoting daytime physical activity and differentiated day-night activity rhythms may also help, though these effects appear secondary to light regularity in this sample. Because our findings concern naturalistic light exposure, behavioural strategies may complement formal light therapy.

Manic symptoms peaked in autumn, consistent with prior prospective evidence of a peak around the autumn equinox [56]. Descriptive light profiles (Figure 1) show a steeper afternoon light decline emerging in autumn; this rapid compression may be more circadian-destabilising than any season’s absolute light level, consistent with evidence that the circadian system is sensitive to rate-of-change rather than its absolute magnitude [57], and may help explain the autumn manic peak. Several factors may explain the absence of mediation. First, our youth sample had predominantly subthreshold manic symptoms, in whom seasonal effects on mania are typically weaker than for depression [5]. Second, YMRS showed significant seasonal variation through the sine component (autumn peak), whereas all actigraphy-derived mediators varied primarily on the cosine (winter-summer axis). This directional mismatch meant that no actigraphy-derived variable was well-positioned to capture the autumn-specific seasonal signal in manic symptoms.

Several limitations should be considered. First, light regularity metrics did not distinguish morning and from evening exposure, which exert opposing effects on health [58, 59] and mental disorders [48]. Second, participants were not followed continuously across all seasons, limiting within-person causal inference. The random intercept structure also assumed uniform individual seasonal sensitivity. Third, medication effects were not modelled and may have altered circadian parameters or light sensitivity [60]. Fourth, Australia’s mild winters mean findings may not generalise to higher-latitude populations. Fifth, pooling cohorts with different diagnostic instruments precluded examination of the contribution of diagnostic subtypes. Finally, moderation analyses by individual characteristics were beyond the present scope.

Future studies should employ light sensors capturing the full outdoor illuminance range and examine time-of-day-specific light regularity. Given emerging evidence of individual differences in photosensitivity—including seasonal variation in light sensitivity [50]— incorporating measures of individual light sensitivity and chronotype would help identify those most vulnerable to seasonal circadian disruption. Finally, intervention studies testing whether promoting greater daytime outdoor light exposure and variability through behavioural coaching, wearable feedback devices, or environmental modifications could direct clinical translation of these findings.

In summary, light exposure variability—reflecting constrained engagement with brighter environments during winter—emerged as the dominant mediator of seasonal mood worsening in Australian youth, with smaller contributions from sleep-activity-circadian patterns. Light exposure variability is a promising, modifiable target for intervention.

## Data Availability

All data produced in the present study are available upon reasonable request to the authors.

## Acknowledgments and Disclosures

This work is supported by a National Health and Medical Research Council (NHMRC) grant (2019206). This work was further supported by NHMRC EL1 Investigator Grants (Grant No. GNT2008196 [to JJC]) and an NHMRC L3 Investigator Grant (Grant No. GNT2016346 [to IBH]).

IBH is the Co-Director, Health and Policy at the Brain and Mind Centre (BMC) University of Sydney, Australia. The BMC operates an early-intervention youth services at Camperdown under contract to headspace. IBH has previously led community-based and pharmaceutical industry-supported (Wyeth, Eli Lilly, Servier, Pfizer, AstraZeneca, Janssen-Cilag) projects focused on the identification and better management of anxiety and depression. He is the Chief Scientific Advisor to, and a 3.2% equity shareholder in, Innowell Pty Ltd., which aims to transform mental health services through the use of innovative technologies. Other authors report no biomedical financial interests or potential conflicts of interest.

## REFERENCES

[1] Mathers CD, Vos ET, Stevenson CE, Begg SJ. The burden of disease and injury in Australia. Bull World Health Organ. 2001;79(11):1076–84.

[2] Patel V, Chisholm D, Parikh R, Charlson FJ, Degenhardt L, Dua T, et al. Addressing the burden of mental, neurological, and substance use disorders: key messages from Disease Control Priorities, 3rd edition. The Lancet. 2016;387(10028):1672–85. 10.1016/S0140-6736(15)00390-6.

[3] Magnusson A, Boivin D. Seasonal Affective Disorder: An Overview. Chronobiology International. 2003;20(2):189–207. 10.1081/CBI-120019310.

[4] Nevarez Flores AG, Bostock EC, Neil AL. Should clinicians and the general population be concerned about seasonal affective disorder in Australia? Med J Aust. 2022;216(10):507–9. 10.5694/mja2.51518.

[5] Estrada-Prat X, Romero S, Borras R, Merranko J, Goldstein T, Hafeman D, et al. Seasonal mood variation in youth and young adults with bipolar spectrum disorder: A longitudinal prospective analysis. Journal of Affective Disorders. 2025;370:159–67. 10.1016/j.jad.2024.10.115.

[6] Chaves MI, Garcês Marques J, Varino F, Siopa C, Duarte AI. Seasonality in bipolar disorder, analysis of an inpatient unit. European Psychiatry. 2025;68(S1):S505–S6. 10.1192/j.eurpsy.2025.1052.

[7] Zhang R, Volkow ND. Seasonality of brain function: role in psychiatric disorders. Translational Psychiatry. 2023;13(1):65. 10.1038/s41398-023-02365-x.

[8] Winthorst WH, Bos EH, Roest AM, de Jonge P. Seasonality of mood and affect in a large general population sample. PLOS ONE. 2020;15(9):e0239033. 10.1371/journal.pone.0239033.

[9] To S, Messias E, Burch L, Chibnall J. Seasonal variation in suicide: age group and summer effects in the United States (2015–2020). BMC Psychiatry. 2024;24(1):856. 10.1186/s12888-024-06309-7.

[10] Christodoulou C, Douzenis A, Papadopoulos FC, Papadopoulou A, Bouras G, Gournellis R, et al. Suicide and seasonality. Acta Psychiatrica Scandinavica. 2012;125(2):127–46. 10.1111/j.1600-0447.2011.01750.x.

[11] Dey D, Lateef HA, Leroux A, Zipunnikov V, Merikangas K. Associations between daily outdoor temperature and subjective real-time ratings of emotional states and sleep in mood disorder subtypes. Journal of Affective Disorders. 2026;397:120918. 10.1016/j.jad.2025.120918.

[12] Crouse JJ, Loblay V, Jorm A, Wong T, de Haan Z, Gorban C, et al. Essential information about chronobiology and chronotherapy for the optimal care of people with bipolar disorders: an international expert consensus. medRxiv. 2025. 10.1101/2025.06.17.25329750.

[13] Di Nicola M, Mazza M, Panaccione I, Moccia L, Giuseppin G, Marano G, et al. Sensitivity to climate and weather changes in euthymic bipolar subjects: association with suicide attempts. Frontiers in psychiatry. 2020;11:95.

[14] Hut RA, Beersma DGM. Evolution of time-keeping mechanisms: early emergence and adaptation to photoperiod. Philosophical Transactions of the Royal Society B: Biological Sciences. 2011;366(1574):2141–54. 10.1098/rstb.2010.0409.

[15] Honma K, Honma S, Kohsaka M, Fukuda N. Seasonal variation in the human circadian rhythm: dissociation between sleep and temperature rhythm. American Journal of Physiology-Regulatory, Integrative and Comparative Physiology. 1992;262(5):R885–R91. 10.1152/ajpregu.1992.262.5.R885.

[16] Meijer JH, Michel S, VanderLeest HT, Rohling JHT. Daily and seasonal adaptation of the circadian clock requires plasticity of the SCN neuronal network. European Journal of Neuroscience. 2010;32(12):2143–51. 10.1111/j.1460-9568.2010.07522.x.

[17] Mattingly SM, Grover T, Martinez GJ, Aledavood T, Robles-Granda P, Nies K, et al. The effects of seasons and weather on sleep patterns measured through longitudinal multimodal sensing. npj Digital Medicine. 2021;4(1):76. 10.1038/s41746-021-00435-2.

[18] Carpenter JS, Crouse JJ, Scott EM, Naismith SL, Wilson C, Scott J, et al. Circadian depression: A mood disorder phenotype. Neurosci Biobehav Rev. 2021;126:79–101. 10.1016/j.neubiorev.2021.02.045.

[19] Duffy JF, Czeisler CA. Effect of Light on Human Circadian Physiology. Sleep Med Clin. 2009;4(2):165–77. 10.1016/j.jsmc.2009.01.004.

[20] Rosenthal SJ, Josephs T, Kovtun O, McCarty R. Seasonal effects on bipolar disorder: A closer look. Neuroscience & Biobehavioral Reviews. 2020;115:199–219. 10.1016/j.neubiorev.2020.05.017.

[21] Scott J, Etain B, Miklowitz D, Crouse JJ, Carpenter J, Marwaha S, et al. A systematic review and meta-analysis of sleep and circadian rhythms disturbances in individuals at high-risk of developing or with early onset of bipolar disorders. Neuroscience & Biobehavioral Reviews. 2022;135:104585. 10.1016/j.neubiorev.2022.104585.

[22] McCarthy MJ, Gottlieb JF, Gonzalez R, McClung CA, Alloy LB, Cain S, et al. Neurobiological and behavioral mechanisms of circadian rhythm disruption in bipolar disorder: A critical multi-disciplinary literature review and agenda for future research from the ISBD task force on chronobiology. Bipolar Disorders. 2022;24(3):232–63. 10.1111/bdi.13165.

[23] Robillard R, Carpenter JS, Rogers NL, Fares S, Grierson AB, Hermens DF, et al. Circadian rhythms and psychiatric profiles in young adults with unipolar depressive disorders. Translational Psychiatry. 2018;8(1):213. 10.1038/s41398-018-0255-y.

[24] Crouse JJ, Carpenter JS, Song YJC, Hockey SJ, Naismith SL, Grunstein RR, et al. Circadian rhythm sleep-wake disturbances and depression in young people: implications for prevention and early intervention. Lancet Psychiatry. 2021;8(9):813–23. 10.1016/s2215-0366(21)00034-1.

[25] Tonini E, Crouse JJ, Shin M, Carpenter JS, Mitchell BL, Byrne EM, et al. Clinical and genetic correlates of a circadian subtype of depression in the Australian Genetics of Depression Study. medRxiv. 2026:2026.02.23.26346917. 10.64898/2026.02.23.26346917.

[26] Parslow RA, Jorm AF, Butterworth P, Jacomb PA, Rodgers B. An examination of seasonality experienced by Australians living in a continental temperate climate zone. Journal of Affective Disorders. 2004;80(2):181–90. 10.1016/S0165-0327(03)00113-7.

[27] Øverland S, Woicik W, Sikora L, Whittaker K, Heli H, Skjelkvåle FS, et al. Seasonality and symptoms of depression: A systematic review of the literature. Epidemiology and Psychiatric Sciences. 2020;29:e31.e31. 10.1017/S2045796019000209.

[28] The Bureau of Meteorology. Average annual & monthly sunshine duration, https://www.bom.gov.au/jsp/ncc/climate_averages/sunshine-hours/index.jsp; 2016.

[29] Ferguson T, Curtis R, Fraysse F, Olds T, Dumuid D, Brown W, et al. The Annual Rhythms in Sleep, Sedentary Behavior, and Physical Activity of Australian Adults: A Prospective Cohort Study. Annals of Behavioral Medicine. 2024;58(4):286–95. 10.1093/abm/kaae007.

[30] Solmi M, Radua J, Olivola M, Croce E, Soardo L, Salazar de Pablo G, et al. Age at onset of mental disorders worldwide: large-scale meta-analysis of 192 epidemiological studies. Molecular Psychiatry. 2022;27(1):281–95. 10.1038/s41380-021-01161-7.

[31] Hickie IB, Scott EM, Cross SP, Iorfino F, Davenport TA, Guastella AJ, et al. Right care, first time: a highly personalised and measurement-based care model to manage youth mental health. Med J Aust. 2019;211 Suppl 9:S3–s46. 10.5694/mja2.50383.

[32] Lee RSC, Hermens DF, Naismith SL, Kaur M, Guastella AJ, Glozier N, et al. Clinical, neurocognitive and demographic factors associated with functional impairment in the Australian Brain and Mind Youth Cohort Study (2008-2016). BMJ Open. 2018;8(12):e022659. 10.1136/bmjopen-2018-022659.

[33] Carpenter JS, Iorfino F, Cross S, Nichles A, Zmicerevska N, Crouse JJ, et al. Cohort profile: the Brain and Mind Centre Optymise cohort: tracking multidimensional outcomes in young people presenting for mental healthcare. BMJ Open. 2020;10(3):e030985. 10.1136/bmjopen-2019-030985.

[34] Nichles A, Zmicerevska N, Song YJC, Wilson C, McHugh C, Hamilton B, et al. Neurobiology Youth Follow-up Study: protocol to establish a longitudinal and prospective research database using multimodal assessments for current and past mental health treatment-seeking young people within an early intervention service. BMJ Open. 2021;11(6):e044731. 10.1136/bmjopen-2020-044731.

[35] Overall JE, Gorham DR. The Brief Psychiatric Rating Scale. Psychological Reports. 1962;10(3):799–812. 10.2466/pr0.1962.10.3.799.

[36] Dazzi F, Shafer A, Lauriola M. Meta-analysis of the Brief Psychiatric Rating Scale – Expanded (BPRS-E) structure and arguments for a new version. Journal of Psychiatric Research. 2016;81:140–51. 10.1016/j.jpsychires.2016.07.001.

[37] Young RC, Biggs JT, Ziegler VE, Meyer DA. A rating scale for mania: reliability, validity and sensitivity. Br J Psychiatry. 1978;133:429–35. 10.1192/bjp.133.5.429.

[38] Rush AJ, Trivedi MH, Ibrahim HM, Carmody TJ, Arnow B, Klein DN, et al. The 16-Item quick inventory of depressive symptomatology (QIDS), clinician rating (QIDS-C), and self-report (QIDS-SR): a psychometric evaluation in patients with chronic major depression. Biological Psychiatry. 2003;54(5):573–83. 10.1016/S0006-3223(02)01866-8.

[39] Goldman HH, Skodol AE, Lave TR. Revising axis V for DSM-IV: a review of measures of social functioning. Am J Psychiatry. 1992;149(9):1148–56. 10.1176/ajp.149.9.1148.

[40] Migueles JH, Rowlands AV, Huber F, Sabia S, van Hees VT. GGIR: A Research Community–Driven Open Source R Package for Generating Physical Activity and Sleep Outcomes From Multi-Day Raw Accelerometer Data. Journal for the Measurement of Physical Behaviour. 2019;2(3):188–96. 10.1123/jmpb.2018-0063.

[41] Guo W, Leroux A, Shou H, Cui L, Kang SJ, Strippoli M-PF, et al. Processing of Accelerometry Data with GGIR in Motor Activity Research Consortium for Health. Journal for the Measurement of Physical Behaviour. 2023;6(1):37–44. 10.1123/jmpb.2022-0018.

[42] Kang SJ, Leroux A, Guo W, Dey D, Strippoli M-PF, Di J, et al. Integrative Modeling of Accelerometry-Derived Sleep, Physical Activity, and Circadian Rhythm Domains With Current or Remitted Major Depression. JAMA Psychiatry. 2024;81(9):911–8. 10.1001/jamapsychiatry.2024.1321.

[43] Hand AJ, Stone JE, Shen L, Vetter C, Cain SW, Bei B, et al. Measuring light regularity: sleep regularity is associated with regularity of light exposure in adolescents. Sleep. 2023;46(8). 10.1093/sleep/zsad001.

[44] R Core Team. A language and environment for statistical computing Vienna, Austria: R foundation for statistical computing. In; 2022.

[45] Bates D, Mächler M, Bolker B, Walker S. Fitting linear mixed-effects models using lme4. Journal of statistical software. 2015;67:1–48.

[46] Shrout PE, Bolger N. Mediation in experimental and nonexperimental studies: new procedures and recommendations. Psychological methods. 2002;7(4):422.

[47] Hayes AF. Beyond Baron and Kenny: Statistical Mediation Analysis in the New Millennium. Communication Monographs. 2009;76(4):408–20. 10.1080/03637750903310360.

[48] Burns AC, Windred DP, Rutter MK, Olivier P, Vetter C, Saxena R, et al. Day and night light exposure are associated with psychiatric disorders: an objective light study in >85,000 people. Nature Mental Health. 2023;1(11):853–62. 10.1038/s44220-023-00135-8.

[49] Adamsson M, Laike T, Morita T. Seasonal Variation in Bright Daylight Exposure, Mood and Behavior among a Group of Office Workers in Sweden. J Circadian Rhythms. 2018;16:2. 10.5334/jcr.153.

[50] Fazlali F, Lazar R, Yahya F, Stefani O, Spitschan M, Cajochen C. Sex and Seasonal Variations in Melatonin Suppression and Alerting Response to Light. J Endocr Soc. 2025;9(12):bvaf155. 10.1210/jendso/bvaf155.

[51] Stone JE, McGlashan EM, Facer-Childs ER, Cain SW, Phillips AJK. Accuracy of the GENEActiv Device for Measuring Light Exposure in Sleep and Circadian Research. Clocks Sleep. 2020;2(2):143–52. 10.3390/clockssleep2020012.

[52] Price LLA, Lyachev A, Khazova M. Optical performance characterization of light-logging actigraphy dosimeters. J Opt Soc Am A. 2017;34(4):545–57. 10.1364/JOSAA.34.000545.

[53] Turrisi TB, Bittel KM, West AB, Hojjatinia S, Hojjatinia S, Mama SK, et al. Seasons, weather, and device-measured movement behaviors: a scoping review from 2006 to 2020. International Journal of Behavioral Nutrition and Physical Activity. 2021;18(1):24. 10.1186/s12966-021-01091-1.

[54] Minaeva O, Booij SH, Lamers F, Antypa N, Schoevers RA, Wichers M, et al. Level and timing of physical activity during normal daily life in depressed and non-depressed individuals. Translational Psychiatry. 2020;10(1):259. 10.1038/s41398-020-00952-w.

[55] Difrancesco S, Penninx B, Riese H, Giltay EJ, Lamers F. The role of depressive symptoms and symptom dimensions in actigraphy-assessed sleep, circadian rhythm, and physical activity. Psychol Med. 2022;52(13):2760–6. 10.1017/s0033291720004870.

[56] Akhter A, Fiedorowicz JG, Zhang T, Potash JB, Cavanaugh J, Solomon DA, et al. Seasonal variation of manic and depressive symptoms in bipolar disorder. Bipolar Disord. 2013;15(4):377–84. 10.1111/bdi.12072.

[57] Parker G, Hadzi-Pavlovic D, Bayes A, Graham R. Relationship between photoperiod and hospital admissions for mania in New South Wales, Australia. Journal of Affective Disorders. 2018;226:72–6. 10.1016/j.jad.2017.09.014.

[58] Windred DP, Burns AC, Rutter MK, Ching Yeung CH, Lane JM, Xiao Q, et al. Personal light exposure patterns and incidence of type 2 diabetes: analysis of 13 million hours of light sensor data and 670,000 person-years of prospective observation. Lancet Reg Health Eur. 2024;42:100943. 10.1016/j.lanepe.2024.100943.

[59] Windred DP, Burns AC, Lane JM, Olivier P, Rutter MK, Saxena R, et al. Brighter nights and darker days predict higher mortality risk: A prospective analysis of personal light exposure in >88,000 individuals. Proc Natl Acad Sci U S A. 2024;121(43):e2405924121. 10.1073/pnas.2405924121.

[60] McGlashan EM, Nandam LS, Vidafar P, Mansfield DR, Rajaratnam SMW, Cain SW. The SSRI citalopram increases the sensitivity of the human circadian system to light in an acute dose. Psychopharmacology (Berl). 2018;235(11):3201–9. 10.1007/s00213-018-5019-0.

