## Supplementary Material for "Seasonal variation in mood and the dynamics of sleep, activity, circadian rhythms, and light"

**Validation of Sine/Cosine Seasonal Model**

To verify that the sine/cosine parameterisation adequately captured seasonal patterns, we compared three models: (1) a null model with random intercepts only; (2) a sine/cosine model adding seasonal terms; and (3) a 12-month categorical model with month as a 12-level factor.

Model comparison used likelihood ratio (LR) tests, information criteria (Akaike Information Criterion [AIC], Bayesian Information Criterion [BIC]), and Pearson correlations between observed and predicted monthly means. LR tests addressed two questions: (a) does seasonality exist (sine/cosine vs null)? and (b) does the 12-month categorical model fit significantly better than sine/cosine?

**Table S1.** Validation of sine/cosine seasonal parameterisation

| **Outcome** | **n** | **r (sine/cos)** | **r (12-month)** | **ΔAIC** | **ΔBIC** | **LR: sine/cos vs null** | **LR: 12-month vs sine/cos** |
| --- | --- | --- | --- | --- | --- | --- | --- |
| QIDS | 527 | 0.683 | 0.873 | -9 | -47 | χ^2^(2) = 6.35,  p = 0.040 | χ^2^(9) = 11.22,  p = 0.429 |
| BPRS Negative | 607 | 0.764 | 0.928 | -14 | -54 | χ^2^(2) = 7.15,  p = 0.027 | χ^2^(9) = 4.33,  p = 0.911 |
| YMRS | 584 | 0.808 | 0.902 | -13 | -52 | χ^2^(2) = 7.30,  p = 0.021 | χ^2^(9) = 8.67,  p = 0.831 |

**Note**: r = Pearson correlation between observed monthly means and model-predicted monthly means. ΔAIC and ΔBIC = sine/cosine minus 12-month categorical; negative values favour the more parsimonious sine/cosine model. LR (sine/cos vs null) tests whether significant seasonality exists (2 df for sine and cosine parameters); LR (12-month vs sine/cos) tests whether the categorical model provides significantly better fit than the smooth sinusoidal model (9 df for 11 month parameters minus 2 sine/cosine parameters).

All three outcomes showed significant seasonality (sine/cosine vs null: all p < 0.05), confirming that seasonal modelling was appropriate. The 12-month categorical model did not significantly improve fit over the sine/cosine model for any outcome (all p > 0.40), indicating that the smooth sinusoidal assumption was adequate. Both AIC and BIC favoured the sine/cosine model (ΔBIC: 47-54 points), supporting its parsimony. Estimated seasonal trajectories with 95% confidence intervals are displayed in Figure 2 (main text). It confirms that the sine/cosine model closely tracked observed monthly means, with symptom peaks occurring during late autumn to winter for depressive and negative symptoms and during autumn for manic symptoms.

**Detailed Actigraphy Data and Processing**

Participants wore GENEActiv accelerometers on the non-dominant wrist for 5-20 consecutive days (median = 12 days). Clinical assessments were administered at the same study visit as actigraphy, typically at the start of each actigraphy recording period. Raw accelerometry data were processed using GGIR (version 3.0.5), an open-source R package for multi-day accelerometer processing [1]. Post-processing followed protocols established for the Mobile Motor Activity Research Consortium for Health (mMARCH) using the mMARCH.AC package (version 2.9.2) [2]. As the GENEActiv device records accelerometry and light exposure simultaneously, light and accelerometry data are inherently concurrent across all observation periods. Light exposure variables were derived from the GGIR Part 5 epoch-level output, which provides minute-level data, rather than from the final GGIR summary tables.

Sleep, circadian rhythm, and physical activity parameters are inter-related but also contain domain-specific information; therefore, the Joint and Individual Variation Explained method (JIVE), a multimodal integrative dimension-reduction technique, was applied to 28 actigraphy-derived variables across three domains (Supplementary Table S2): sleep (7 variables), physical activity (7 variables), and circadian rhythm (14 variables) [3]. The analysis decomposed total variance into joint (shared across domains), individual (domain-specific), and residual components. Loadings and proportional variance explained were examined to support interpretability and the detailed component structure is reported in the Results and Supplementary Table S3.

Light parameters were not included in the JIVE dimension reduction, as they represent a different source of circadian input to the accelerometry-based data. Raw wrist-measured light data are presented descriptively to characterise the ambient light environment across seasons (Figure 1); however, per-minute light values reflect a combination of environmental availability, individual behaviour, and sensor artefact, and are not suitable as individual-level predictors. Accordingly, Principal Component Analysis (PCA) was conducted separately to characterise dimensions of light exposure patterns. Following a previous study [4], light exposure was characterised using three metric types across seven lux thresholds (10, 20, 50, 100, 300, 500, 1,000 lux*): Light Regularity Index (LRI)*, measuring day-to-day consistency of light exposure timing; *mean light timing (MLiT)*, representing the average clock time of light exposure above each threshold; and *photoperiod interval*, capturing the duration between first and last light exposure above each threshold. After quality control (see Supplementary Materials), complete light data across all 21 variables were available for 491 observation periods from 254 participants (67.0%). Light exposure measurements are driven by highly time- and context-specific environmental factors (e.g., weather conditions, location, and individual behaviour) that were not captured by the study covariates. As a result, the missingness mechanism is unlikely to satisfy the missing at random assumption required for standard imputation methods. To avoid introducing model-based bias, analyses were therefore restricted to complete cases.

Given the inherent within- and between-participant variability in light exposure, PCA with oblimin rotation was conducted on the complete cases sample to identify latent dimensions of light exposure patterns. To address potential concerns about the inclusion of repeated observations in PCA, sensitivity analyses were used to evaluate the robustness of the solution: comparing all available observations (n = 491) versus first observations only (n = 253) yielded near-identical factor structures (Tucker’s congruence coefficients 0.995-1.000). Additionally, excluding 1,000 lux threshold variables (which had 33% missingness) retained the four-component structure with consistent loadings and interpretations (Tucker’s congruence coefficients 0.935-0.994; see Supplementary Materials).

**Joint and Individual Variation Explained (JIVE) Analysis**

**Table S2.** Variables included in the Joint and Individual Variation Explained (JIVE) Analysis

| Domain | Variables |
| --- | --- |
| Sleep | Sleep onset |
|  | Sleep wakeup |
|  | Sleep midpoint |
|  | Sleep duration |
|  | Sleep efficiency |
|  | Number of times awake during the night for at least 5 minutes (NWB) |
|  | Number of blocks of night sleep within sleep period (NSB) |
| Physical activity | Total accelerometry count (TAC) |
|  | Total log-transformed accelerometry count (TLAC) |
|  | Total sedentary time (TST) |
|  | Total number of minutes spent in light intensity physical activity (50 < activity count < 100) (LiPA) |
|  | Total number of minutes spent in moderate to vigorous physical activity (activity count > 100) (MVPA) |
|  | Sedentary to active transition probability (SATP) |
|  | Active to sedentary transition probability (ASTP) |
| Circadian rhythms | fPC1 (1st principal component score from functional principal component analysis) |
|  | fPC2 (2nd principal component score from functional principal component analysis) |
|  | fPC3 (3rd principal component score from functional principal component analysis) |
|  | fPC4 (4th principal component score from functional principal component analysis) |
|  | Relative amplitude (RA) |
|  | Intra-daily variability (IV) |
|  | Inter-day stability (IS) |
|  | Midline-estimating statistic of rhythm, a rhythm adjusted mean (MESOR) |
|  | Amplitude, a measure of variability around the mean (Amp) |
|  | Acrophase, a measure of timing of the peak value of cycle (Acro) |
|  | Acceleration value of the most active 10 hours (M10) |
|  | Acceleration value of the least active 5 hours (L5) |
|  | Timing (circular mean) of most active 10 hours (M10 Time) |
|  | Timing (circular mean) of least active 5 hours (L5 Time) |

**Note:** JIVE analysis follows the methodology in a previous study[3].

**Figure S1.** JIVE variance decomposition across domains

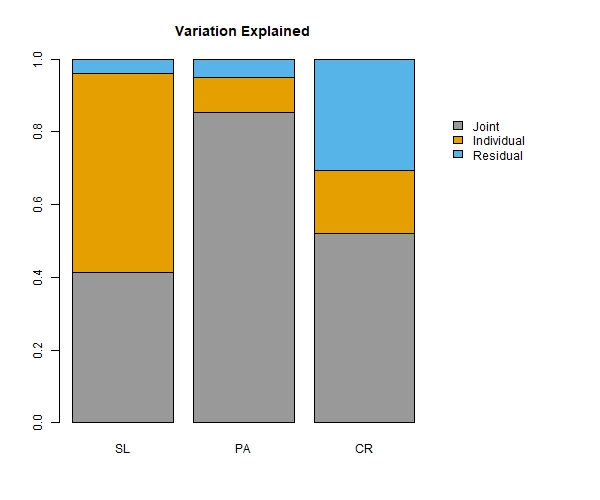

Joint components accounted for 41.4% of the total variation in the sleep domain, 85.4% of the total variation in the physical activity domain, and 52.2% of the total variation in the circadian rhythm domain. These high levels of joint variation indicate the substantial overlap across these three domains. Individual components explained 54.5%, 9.5%, and 17.3% of the total variation in sleep, physical activity, and circadian rhythm domains, respectively. Only 4.1%, 5.1%, and 30.5% of the total variation in each domain remained unexplained by the estimated JIVE model.

**Table S3.** JIVE component loadings across joint and individual domains

| Variables | V1 | V2 | V3 |
| --- | --- | --- | --- |
| **Joint** | | | |
| Sleep onset | -0.00067 | **0.493647** | -0.118 |
| Sleep wakeup | -0.09967 | **0.42428** | 0.180001 |
| Sleep midpoint | -0.05742 | **0.509182** | 0.033409 |
| Sleep duration | -0.07958 | -0.10974 | **0.394894** |
| Sleep efficiency | 0.04689 | -0.06626 | 0.170493 |
| NSB | -0.0557 | 0.021788 | 0.007462 |
| NWB | -0.06419 | 0.026195 | -0.03721 |
| TAC | **0.341336** | 0.039743 | -0.06057 |
| TLAC | **0.315006** | 0.058108 | -0.12831 |
| TST | **-0.34116** | -0.04038 | -0.03328 |
| LiPA | **0.303633** | 0.041542 | 0.090317 |
| MVPA | **0.327531** | 0.035911 | -0.05004 |
| SATP | **0.304534** | 0.034673 | 0.018976 |
| ASTP | **-0.30135** | -0.02947 | -0.05909 |
| RA | 0.207288 | 0.003525 | 0.131654 |
| IV | -0.14287 | -0.03384 | **-0.25616** |
| IS | 0.09566 | -0.03462 | 0.234846 |
| Mesor | 0.241344 | 0.0281 | -0.04283 |
| Amp | 0.229157 | 0.023789 | 0.054533 |
| Acro | 0.042345 | -0.30189 | -0.0237 |
| L5 Time | -0.04469 | **0.311909** | 0.04657 |
| M10 Time | -0.07039 | **0.276886** | 0.04182 |
| L5 | 0.0412 | 0.084797 | **-0.27133** |
| M10 | 0.238322 | 0.029118 | 0.0061 |
| fPC1 | -0.05834 | 0.033702 | **0.439252** |
| fPC2 | -0.03484 | -0.09152 | 0.091156 |
| fPC3 | -0.01147 | 0.037063 | 0.078216 |
| fPC4 | -0.02421 | -0.00654 | **-0.54949** |
| **Sleep** |  |  |  |
| Sleep onset | -0.14206 | 0.138556 | NA |
| Sleep wakeup | 0.125306 | **-0.40084** | NA |
| Sleep midpoint | -0.00577 | -0.14488 | NA |
| Sleep duration | 0.086922 | **-0.80045** | NA |
| Sleep efficiency | **-0.52212** | **-0.37204** | NA |
| NSB | **0.578389** | -0.13598 | NA |
| NWB | **0.591089** | 0.039003 | NA |
| **Physical activity** | | | |
| TAC | -0.05794 | NA | NA |
| TLAC | **0.259001** | NA | NA |
| TST | -0.14304 | NA | NA |
| LiPA | **0.504355** | NA | NA |
| MVPA | **-0.31223** | NA | NA |
| SATP | **0.554243** | NA | NA |
| ASTP | **0.500056** | NA | NA |
| **Circadian Rhythm** |  |  |  |
| RA | 0.060614 | **0.307607** | NA |
| IV | -0.18276 | **-0.25502** | NA |
| IS | **0.257168** | **0.27541** | NA |
| Mesor | -0.02921 | -0.01925 | NA |
| Amp | 0.042547 | 0.063341 | NA |
| Acro | 0.199365 | -0.04395 | NA |
| L5 Time | -0.06697 | -0.0061 | NA |
| M10 Time | -0.24582 | -0.03613 | NA |
| L5 | -0.11319 | -0.60632 | NA |
| M10 | 0.001818 | 0.010382 | NA |
| fPC1 | **-0.43858** | 0.157205 | NA |
| fPC2 | **0.311942** | -0.3589 | NA |
| fPC3 | **0.697434** | -0.02293 | NA |
| fPC4 | -0.02395 | **0.484828** | NA |

**Note:** Loadings with absolute values >0.25 indicate meaningful contributions to the component.

**Figure S2.** Functional principal component analysis (fPCA) of 24-hour activity profiles

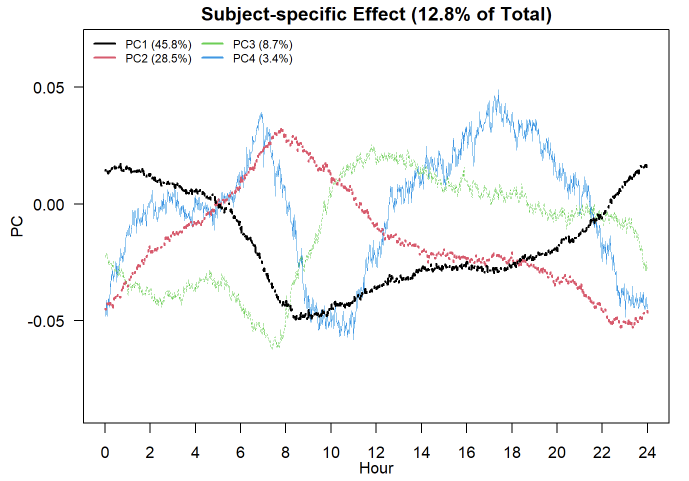

**Principal Component Analysis (PCA) of light exposure**

**Light exposure variables:** Twenty-one light exposure variables were included to comprehensively characterise light patterns[4]: Light Regularity Index (LRI); mean light timing variables (milt); photoperiod intervals, each calculated at 10, 20, 50, 100, 300, 500, 1,000 lux thresholds.

**Quality Controls:** Light data underwent systematic quality control: (1) invalid epochs were removed based on sleep/wake state classification; (2) bins where neighbouring bins showed intensity 10 times higher (indicating sensor artefact) were removed; (3) days with >80% of data below 10 lux, >75% above 6000 lux, or >80% identical values (sensor malfunction) were excluded; and (4) participants required at least one weekend day and two weekdays of valid data. After quality control, 491 observations from 254 participants had complete light data. Missing data increased with lux threshold (9.3% at 10-300 lux; 33.0% at 1,000 lux; all other thresholds had 9-20% missing).

**PCA component selection:** PCA suitability was confirmed (Kaiser-Meyer-Olkin = 0.79; Bartlett’s test: χ^2^(210) = 19,268, p < 0.001). Multiple criteria converged on retaining four components explaining 84.6% of variance: parallel analysis, Kaiser criterion (eigenvalues: 7.9, 4.4, 3.3, 2.2, 0.9...), and scree plot inspection (Figure S3).

**Table S4.** Light exposure PCA component loadings (N = 491)

| **Variable** | **PC1** | **PC2** | **PC3** | **PC4** |
| --- | --- | --- | --- | --- |
| LRI_10 | 0.03 | -0.13 | **0.83** | -0.01 |
| LRI_20 | 0.03 | -0.09 | **0.93** | -0.05 |
| LRI_50 | 0.03 | 0 | **0.97** | 0.01 |
| LRI_100 | -0.02 | 0.04 | **0.9** | 0.26 |
| LRI_300 | -0.04 | -0.02 | 0.36 | **0.88** |
| LRI_500 | -0.02 | -0.04 | 0.05 | **0.96** |
| LRI_1000 | 0 | -0.02 | -0.17 | **0.88** |
| interval_10_comb | 0.14 | **0.71** | -0.34 | 0.21 |
| interval_20_comb | 0.13 | **0.74** | -0.33 | 0.19 |
| interval_50_comb | 0.11 | **0.78** | -0.3 | 0.16 |
| interval_100_comb | 0.08 | **0.86** | -0.2 | 0.08 |
| interval_300_comb | -0.03 | **0.94** | 0.1 | -0.11 |
| interval_500_comb | -0.04 | **0.89** | 0.23 | -0.23 |
| interval_1000_comb | -0.09 | **0.73** | 0.34 | -0.34 |
| mlit_10_comb | **0.88** | 0 | 0.12 | 0.16 |
| mlit_20_comb | **0.93** | 0 | 0.07 | 0.11 |
| mlit_50_comb | **0.96** | -0.01 | 0.06 | 0.05 |
| mlit_100_comb | **0.96** | -0.02 | 0.02 | -0.03 |
| mlit_300_comb | **0.93** | 0.02 | -0.04 | -0.13 |
| mlit_500_comb | **0.86** | 0.03 | -0.06 | -0.16 |
| mlit_1000_comb | **0.76** | 0.03 | -0.08 | -0.18 |
| **Eigenvalue** | 7.86 | 4.41 | 3.27 | 2.23 |
| **Variance (%)** | 37.4 | 21 | 15.6 | 10.6 |
| **Cumulative (%)** | 37.4 | 58.5 | 74 | 84.6 |

**Note**: Principal component analysis with oblimin rotation. PC1 = timing of light exposure (MLiT); PC2 = duration/photoperiod (interval); PC3 = regularity of dim-moderate light (LRI 10-100 lux); PC4 = regularity of bright ambient light (LRI 300-1,000 lux). Loadings > 0.50 indicate primary contributions.

**Figure S3.** A parallel analysis and scree plot for light exposure PCA.

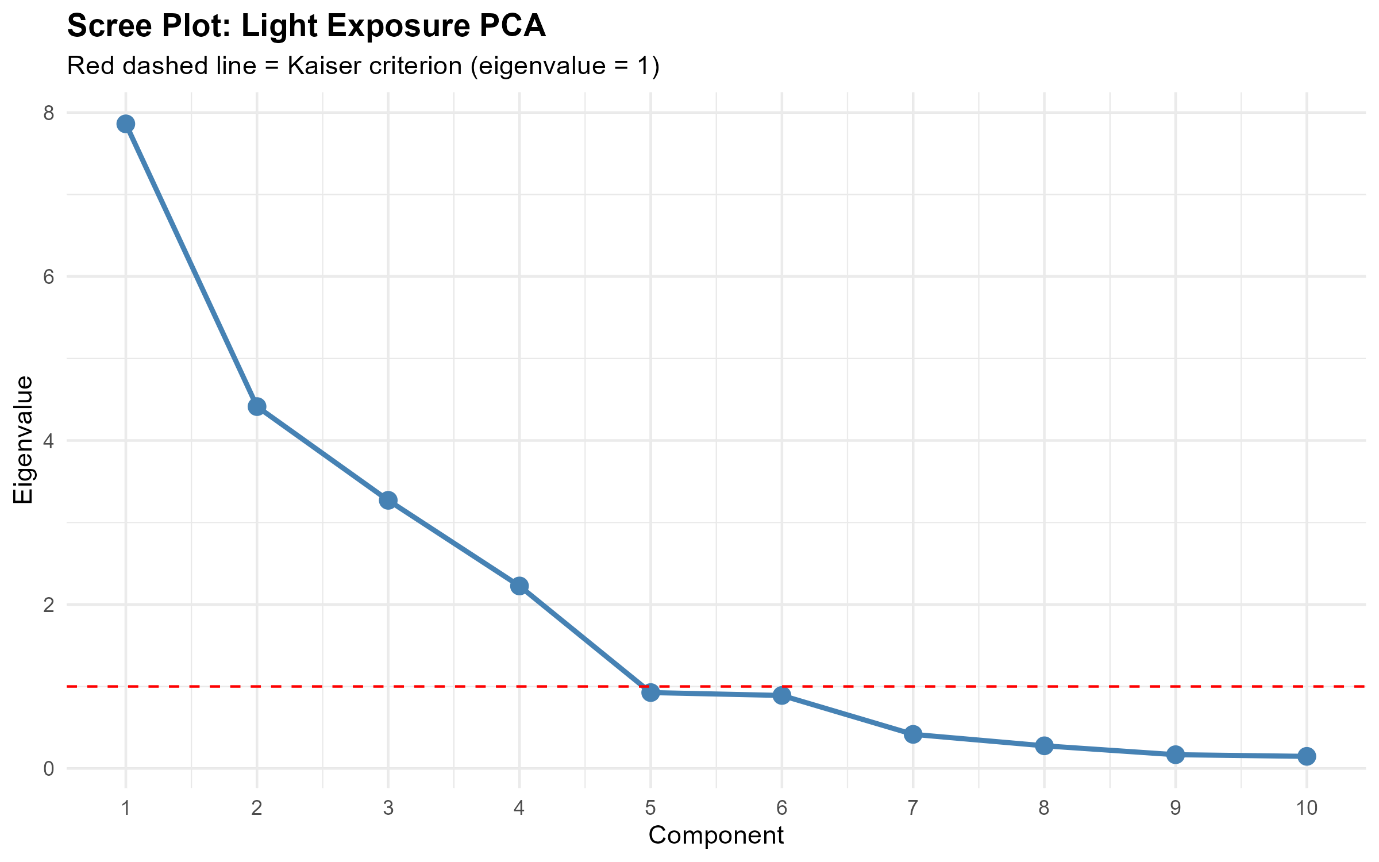

**Note**: Scree plot showing observed eigenvalues (solid line) versus 95th percentile of eigenvalues from parallel analysis with random data (dashed line). Four components exceeded both the Kaiser criterion (eigenvalue >1) and parallel analysis threshold.

**Principal Component Analysis (PCA) of light exposure – Sensitivity Analyses**

**PCA robustness to repeated observations.** A sensitivity analysis compared PCA conducted on all available observations (N = 491) versus first observations only (N = 253). Tucker’s congruence coefficients were 1.000, 0.999, 0.999, and 0.995 for PC1 through PC4 respectively, indicating virtually identical factor structures. Variance explained was also comparable (82.5% for all observations vs 82.4% for first observations only). These results support the robustness of derived components to the inclusion of repeated measurements.

**Table S5.** Tucker’s congruence coefficients comparing PCA solutions: all observations versus first observations only

| **Component** | **Congruence coefficient** |
| --- | --- |
| PC1 (Timing) | 1.000 |
| PC2 (Duration) | 0.999 |
| PC3 (LRI dim) | 0.999 |
| PC4 (LRI bright) | 0.995 |

**Note**: Tucker’s congruence coefficients >0.95 indicate highly similar factor structures between PCA solutions. All observations: N = 491; First observations only: N = 253.

**Impact of 1000 lux threshold variables.** Given the higher missingness (33%) at the 1000 lux threshold, we conducted sensitivity analyses comparing PCA solutions with all 21 light variables versus 18 variables (excluding LRI_1000, interval_1000, and MLiT_1000). Excluding 1,000 lux variables increased the available sample to 586 observations.

The four-component structure remained stable (Table S6). Tucker’s congruence coefficients comparing the 18 common variables across solutions were 0.994, 0.964, 0.961, and 0.935 for PC1 through PC4 respectively, indicating excellent correspondence for timing, duration, and dim light regularity components, and fair correspondence for bright light regularity. The slightly lower congruence for PC4 was expected, as this component captured LRI 300-1000 lux in the main analysis but only LRI 300-500 lux when 1,000 lux variables were excluded. Critically, the loading patterns and component interpretations were consistent across both analyses, supporting the robustness of derived components.

**Table S6.** Light exposure PCA component loadings without 1,000 lux variables (N = 586)

| **Variable** | **PC1** | **PC2** | **PC3** | **PC4** |
| --- | --- | --- | --- | --- |
| LRI_10 |  |  | 0.832 |  |
| LRI_20 |  |  | 0.913 |  |
| LRI_50 |  |  | 0.964 |  |
| LRI_100 |  |  | 0.901 |  |
| LRI_300 |  |  |  | 0.958 |
| LRI_500 |  |  |  | 0.877 |
| interval_10_comb |  | 0.917 |  |  |
| interval_20_comb |  | 0.94 |  |  |
| interval_50_comb |  | 0.956 |  |  |
| interval_100_comb |  | 0.926 |  |  |
| interval_300_comb |  | 0.736 |  |  |
| interval_500_comb |  | 0.511 |  | -0.556 |
| mlit_10_comb | 0.912 |  |  |  |
| mlit_20_comb | 0.959 |  |  |  |
| mlit_50_comb | 0.975 |  |  |  |
| mlit_100_comb | 0.958 |  |  |  |
| mlit_300_comb | 0.851 |  |  |  |
| mlit_500_comb | 0.699 |  |  |  |
| **Eigenvalue** | 6.84 | 3.82 | 3.60 | 1.53 |
| **Variance (%)** | 38.0 | 21.2 | 20.0 | 8.5 |
| **Cumulative (%)** | 38.0 | 59.2 | 79.2 | 87.7 |

**Note**: Loadings <0.50 suppressed for clarity. Component interpretations match main analysis: PC1 = timing of light exposure; PC2 = duration/photoperiod; PC3 = regularity of dim-moderate light (10-100 lux); PC4 = regularity of bright ambient light (300-500 lux).

**Detailed Statistical Analysis**

All analyses were conducted in R version 4.5.0[5] using the lme4 package for linear mixed-effects models[6]. This study adopted an exploratory approach; therefore, no corrections for multiple comparisons were applied. All models included age and sex as covariates and a random intercept for participant to account for repeated observation periods from the same individual.

**Aim 1:** Seasonal effects were examined for seven clinical outcomes (QIDS total score; YMRS total score; BPRS four subscales [affect, positive, negative, activation]; SOFAS score) using linear mixed-effects models with both sine and cosine seasonal terms entered simultaneously:

$$Y_{ij}=\beta_{0}+\beta_{1} sin\left( \frac{2\pi s_{ij}}{365.25} \right)+\beta_{2} cos\left( \frac{2\pi s_{ij}}{365.25} \right)+\beta_{3}Age_{i} +\beta_{4} Sex_{i} + u_{i} +\varepsilon_{ij}$$

Where *i* indexes participant, *j* indexes observation period, $s_{ij}$ is the day-of-year of the actigraphy start date (which coincided with the clinical assessment date). The participant-level random intercept $u_{i} \sim N(0, \sigma_{u}^{2})$ captures between-person differences in average symptom levels, and $\varepsilon_{ij} \sim N(0, \sigma^{2}\varepsilon)$ is the residual error. Evidence of seasonality was defined as p < 0.05 for either seasonal component. Estimated seasonal trajectories for significant outcomes are displayed alongside observed monthly means in Figure 2 (see Results).

**Aim 2:** For those outcomes demonstrating significant seasonality (see Aim 1), mediation analyses were subsequently conducted to examine whether sleep-activity-circadian or light exposure variables mediated seasonal effects on the clinical outcome. As clinical assessments were conducted at the commencement of each actigraphy recording period (with actigraphy-derived mediators reflecting behaviour over the subsequent days) strict temporal precedence cannot be established; findings should therefore be interpreted as reflecting concurrent seasonal patterning. Twelve candidate mediators (8 JIVE-derived sleep-activity-circadian components and 4 PCA-derived light components) were tested without pre-selection, consistent with recommendations to avoid biased screening based on individual paths in mediation models[7, 8]. All variables were standardised prior to mediation analysis to enable comparison of effect sizes across mediators. Seasonal sine and cosine terms were divided by √0.5 (their theoretical standard deviation under uniform distribution). Mediator and outcome variables were z-score transformed. Indirect effects (a x b) were first screened using Sobel test. Mediators showing preliminary evidence of mediation (Sobel p < 0.10) were subsequently tested using cluster bootstrap resampling (1,000 iterations), with participants as the resampling unit to preserve the within-person correlation structure. Bootstrap 95% confidence intervals and p-values were derived from the resampled distributions of indirect effects.

Missing data arose from three sources. First, QIDS was collected only in the Neurobiology cohort, resulting in 206 missing observations (28.1%). Second, BPRS and YMRS had item-level non-response in a subset of assessments (BPRS Negative: 126 missing, 17.2%; YMRS: 149 missing, 20.3%). Third, light PCA scores were unavailable for 242 observations (33.0%), primarily because the 1,000 lux threshold variables were undefined for participants who did not accumulate sufficient exposure at that intensity, rather than reflecting recording failure. All analyses used listwise deletion, with the analytic sample varying by outcome and mediator as a result. Observation periods with and without complete light data for PCA are compared in Supplementary Table S7; those without complete light data were younger (mean 22.4 vs 26.3 years, p < 0.001) but did not differ meaningfully in clinical severity—although BPRS Negative reached statistical significance (p = 0.001), medians were identical. This suggests exclusion reflects data availability rather than systematic clinical selection bias.

**Table S7.** Characteristics of observation periods included versus excluded from light exposure analyses

| Group | Excluded (n=242) | Included (n=491) | p-value |
| --- | --- | --- | --- |
| Age | 22.4±4.7 | 26.3±5.6 | <0.001 |
| Female (%) | 65.3% | 61.1% | 0.307 |
| QIDS | 10.8±5.4 | 9.4±4.9 | 0.124 |
| BPRS Negative | 7.4±2.4 | 6.6±1.2 | 0.001 |
| YMRS | 2.1±3.5 | 1.2±1.7 | 0.104 |

**Note:** Data are presented as mean±SD or percentage. The unit of analysis is the observation period (n=733 total). Inclusion in light exposure analyses required complete data across all 21 light variables; missingness was primarily driven by the 1,000 lux threshold variables (33.0% missing), which were undefined for observation periods where participants did not accumulate sufficient exposure at that intensity. Group differences were tested using Wilcoxon rank-sum tests for continuous variables and chi-square test for sex. QIDS = Quick Inventory of Depressive Symptomatology (Neurobiology cohort only); BPRS = Brief Psychiatric Rating Scale; YMRS = Young Mania Rating Scale.

Table S7 compares characteristics of observation periods included versus excluded from the light exposure analyses. Excluded observation periods were significantly younger (mean 22.4±4.7 vs 26.3±5.6 years, p<0.001). Sex (p=0.307), depressive symptoms (p=0.124), and manic symptoms (p=0.104) did not differ between groups. Although BPRS Negative scores reached statistical significance (p=0.001), the medians were identical between groups (6.0 [IQR 6.0–7.0]) and the mean difference (7.4 vs 6.6) likely reflects skewness from high-scoring outliers in the excluded group rather than a systematic clinical difference. Taken together, exclusion from light analyses appears to reflect differences in data availability rather than systematic selection by clinical severity

**Table S8.** Participant characteristics by visit

|  | Overall | Visit 1 | Visit 2 | Visit 3+ |
| --- | --- | --- | --- | --- |
| N | 733 | 422 | 174 | 137 |
| Age | 25.0±5.7 | 24.3±5.5 | 25.8±5.9 | 26.3±5.5 |
| Female (%) | 62.5% | 63.3% | 62.6% | 59.9% |
| QIDS | 9.5±5.0 | 10.4 ±5.1 | 8.9±4.7 | 8.3 ±4.7 |
| BPRS Depressive | 12.4±4.4 | 12.8 ±4.4 | 12.3±4.5 | 11.6±4.2 |
| BPRS Positive | 6.7±1.6 | 7.0±1.9 | 6.5±1.1 | 6.3±0.8 |
| BPRS Negative | 6.7±1.6 | 6.8±1.7 | 6.6±1.4 | 6.6±1.4 |
| BPRS Manic | 8.5±1.9 | 8.7±2.0 | 8.4±1.7 | 8.4±1.8 |
| YMRS | 1.4±2.2 | 1.5±2.1 | 1.3±2.0 | 1.0±2.3 |
| SOFAS | 65.8±14.4 | 64.9±14.2 | 66.4±13.8 | 67.5±15.6 |

**Note:** Data are presented as mean±SD or percentage. Participants contributing multiple visits appear in more than one column. BPRS = Brief Psychiatric Rating Scale; QIDS = Quick Inventory of Depressive Symptomatology (Neurobiology cohort only); YMRS = Young Mania Rating Scale; SOFAS = Social and Occupational Functioning Assessment Scale.

**Mediation Analysis Results**

**Table S9.** Mediation analysis results for all candidate mediators for significant outcomes

| **Mediator** | **N**  **(obs)** | **N**  **(sample)** | **Path a**  **(SE)** | **p**  **(Path**  **a)** | **Path b**  **(SE)** | **P**  **(Path**  **b)** | **Total**  **effect (c,**  **SE)** | **p**  **(Total**  **effect)** | **Direct**  **effect (c',**  **SE)** | **p**  **(Direct**  **effect)** | **Indirect**  **(Sobel)** | **p**  **(Sobel)** | **Indirect**  **(Bootstrap)** | **p**  **(Bootstrap)** | **%**  **mediated** |
| --- | --- | --- | --- | --- | --- | --- | --- | --- | --- | --- | --- | --- | --- | --- | --- |
| **Depression (QIDS)** | | | | | | | | | | | | | | | |
| JScore_CR_1 | 527 | 266 | -0.09 (0.045) | 0.048 | 0.065 (0.041) | 0.112 | -0.093 (0.041) | 0.025 | -0.087 (0.041) | 0.034 | -0.0058 [-0.015, 0.003] | 0.215 | — | — | — |
| JScore_CR_2 | 527 | 266 | 0.027 (0.043) | 0.530 | -0.021 (0.042) | 0.612 | -0.093 (0.041) | 0.025 | -0.092 (0.041) | 0.026 | -0.0006 [-0.003, 0.002] | 0.693 | — | — | — |
| **JScore_Joint_1** | **527** | **266** | **0.084 (0.029)** | **0.005** | **-0.103 (0.046)** | **0.026** | **-0.093 (0.041)** | **0.025** | **-0.086 (0.041)** | **0.038** | **-0.0086 [-0.018, 0.001]** | **0.079** | **-0.0086 [-0.021, -0.001]** | **0.032** | **9.3** |
| JScore_Joint_2 | 527 | 266 | 0.052 (0.036) | 0.152 | 0.091 (0.045) | 0.042 | -0.093 (0.041) | 0.025 | -0.095 (0.041) | 0.020 | 0.0048 [-0.003, 0.013] | 0.241 | — | — | — |
| JScore_Joint_3 | 527 | 266 | -0.131 (0.044) | 0.003 | 0.051 (0.042) | 0.228 | -0.093 (0.041) | 0.025 | -0.086 (0.041) | 0.038 | -0.0066 [-0.018, 0.005] | 0.264 | — | — | — |
| JScore_PA_1 | 527 | 266 | 0.061 (0.034) | 0.077 | 0.154 (0.046) | 0.001 | -0.093 (0.041) | 0.025 | -0.101 (0.041) | 0.014 | 0.0094 [-0.002, 0.021] | 0.116 | — | — | — |
| JScore_SL_1 | 527 | 266 | 0.006 (0.034) | 0.862 | 0.03 (0.046) | 0.514 | -0.093 (0.041) | 0.025 | -0.093 (0.041) | 0.025 | 0.0002 [-0.002, 0.002] | 0.866 | — | — | — |
| JScore_SL_2 | 527 | 266 | 0.116 (0.036) | 0.001 | 0.083 (0.047) | 0.075 | -0.093 (0.041) | 0.025 | -0.102 (0.041) | 0.014 | 0.0096 [-0.002, 0.022] | 0.119 | — | — | — |
| Light_PC1 | 484 | 247 | 0.313 (0.042) | 0.000 | 0.088 (0.046) | 0.056 | -0.105 (0.043) | 0.015 | -0.132 (0.045) | 0.003 | 0.0275 [-0.002, 0.056] | 0.063 | 0.0275 [-0.002, 0.059] | 0.072 | -29.7 |
| Light_PC2 | 484 | 247 | 0.134 (0.04) | 0.001 | 0.014 (0.047) | 0.762 | -0.105 (0.043) | 0.015 | -0.106 (0.043) | 0.014 | 0.0019 [-0.010, 0.014] | 0.763 | — | — | — |
| Light_PC3 | 484 | 247 | 0.009 (0.038) | 0.811 | -0.02 (0.047) | 0.671 | -0.105 (0.043) | 0.015 | -0.104 (0.043) | 0.015 | -0.0002 [-0.002, 0.002] | 0.835 | — | — | — |
| **Light_PC4** | **484** | **247** | **-0.407 (0.039)** | **0.000** | **0.142 (0.048)** | **0.003** | **-0.105 (0.043)** | **0.015** | **-0.047 (0.047)** | **0.316** | **-0.0577 [-0.098, -0.018]** | **0.005** | **-0.0577 [-0.103, -0.016]** | **0.006** | **62.1** |
| **Mania (YMRS)** | | | | | | | | | | | | | | | |
| JScore_CR_1 | 584 | 314 | 0.06 (0.038) | 0.111 | -0.041 (0.037) | 0.272 | 0.08 (0.033) | 0.015 | 0.082 (0.033) | 0.012 | -0.0025 [-0.008, 0.003] | 0.365 | — | — | — |
| JScore_CR_2 | 584 | 314 | -0.039 (0.036) | 0.278 | -0.024 (0.039) | 0.537 | 0.08 (0.033) | 0.015 | 0.079 (0.033) | 0.016 | 0.0009 [-0.002, 0.004] | 0.591 | — | — | — |
| JScore_Joint_1 | 584 | 314 | -0.027 (0.024) | 0.267 | 0.072 (0.044) | 0.101 | 0.08 (0.033) | 0.015 | 0.081 (0.033) | 0.013 | -0.0019 [-0.006, 0.002] | 0.357 | — | — | — |
| JScore_Joint_2 | 584 | 314 | -0.017 (0.031) | 0.581 | 0.013 (0.041) | 0.753 | 0.08 (0.033) | 0.015 | 0.08 (0.033) | 0.014 | -0.0002 [-0.002, 0.001] | 0.785 | — | — | — |
| JScore_Joint_3 | 584 | 314 | 0.002 (0.037) | 0.964 | 0.051 (0.038) | 0.177 | 0.08 (0.033) | 0.015 | 0.08 (0.033) | 0.014 | 0.0001 [-0.004, 0.004] | 0.964 | — | — | — |
| JScore_PA_1 | 584 | 314 | 0.004 (0.029) | 0.893 | 0.023 (0.043) | 0.596 | 0.08 (0.033) | 0.015 | 0.08 (0.033) | 0.015 | 0.0001 [-0.001, 0.001] | 0.896 | — | — | — |
| JScore_SL_1 | 584 | 314 | -0.003 (0.028) | 0.902 | -0.05 (0.043) | 0.241 | 0.08 (0.033) | 0.015 | 0.079 (0.033) | 0.016 | 0.0002 [-0.003, 0.003] | 0.902 | — | — | — |
| JScore_SL_2 | 584 | 314 | 0.001 (0.031) | 0.968 | -0.007 (0.042) | 0.860 | 0.08 (0.033) | 0.015 | 0.08 (0.033) | 0.015 | -0.0000 [-0.000, 0.000] | 0.969 | — | — | — |
| Light_PC1 | 483 | 246 | -0.04 (0.039) | 0.302 | -0.037 (0.037) | 0.316 | 0.063 (0.034) | 0.061 | 0.062 (0.034) | 0.069 | 0.0015 [-0.003, 0.006] | 0.471 | — | — | — |
| Light_PC2 | 483 | 246 | -0.039 (0.035) | 0.268 | 0.048 (0.038) | 0.211 | 0.063 (0.034) | 0.061 | 0.065 (0.034) | 0.054 | -0.0019 [-0.006, 0.003] | 0.406 | — | — | — |
| Light_PC3 | 483 | 246 | -0.004 (0.033) | 0.904 | -0.069 (0.038) | 0.072 | 0.063 (0.034) | 0.061 | 0.063 (0.034) | 0.061 | 0.0003 [-0.004, 0.005] | 0.905 | — | — | — |
| Light_PC4 | 483 | 246 | 0.034 (0.038) | 0.378 | -0.011 (0.037) | 0.768 | 0.063 (0.034) | 0.061 | 0.064 (0.034) | 0.060 | -0.0004 [-0.003, 0.002] | 0.779 | — | — | — |
| **Negative Symptoms (BPRS)** | | | | | | | | | | | | | | | |
| JScore_CR_1 | 607 | 329 | -0.088 (0.043) | 0.042 | 0.021 (0.036) | 0.565 | -0.08 (0.037) | 0.031 | -0.078 (0.037) | 0.035 | -0.0018 [-0.008, 0.005] | 0.579 | — | — | — |
| JScore_CR_2 | 607 | 329 | 0.028 (0.041) | 0.490 | -0.049 (0.039) | 0.202 | -0.08 (0.037) | 0.031 | -0.078 (0.037) | 0.035 | -0.0014 [-0.006, 0.003] | 0.543 | — | — | — |
| JScore_Joint_1 | 607 | 329 | 0.067 (0.028) | 0.017 | -0.075 (0.046) | 0.103 | -0.08 (0.037) | 0.031 | -0.075 (0.037) | 0.042 | -0.0050 [-0.012, 0.002] | 0.177 | — | — | — |
| JScore_Joint_2 | 607 | 329 | 0.05 (0.035) | 0.153 | 0.026 (0.042) | 0.530 | -0.08 (0.037) | 0.031 | -0.081 (0.037) | 0.029 | 0.0013 [-0.003, 0.006] | 0.565 | — | — | — |
| **JScore_Joint_3** | **607** | **329** | **-0.125 (0.042)** | **0.003** | **0.12 (0.037)** | **0.001** | **-0.08 (0.037)** | **0.031** | **-0.066 (0.037)** | **0.073** | **-0.0150 [-0.028, -0.002]** | **0.029** | **-0.0150 [-0.031, -0.003]** | **<0.001** | **18.7** |
| JScore_PA_1 | 607 | 329 | 0.072 (0.033) | 0.030 | 0 (0.043) | 0.999 | -0.08 (0.037) | 0.031 | -0.08 (0.037) | 0.032 | -0.0000 [-0.006, 0.006] | 0.999 | — | — | — |
| JScore_SL_1 | 607 | 329 | 0.006 (0.032) | 0.863 | -0.016 (0.044) | 0.718 | -0.08 (0.037) | 0.031 | -0.08 (0.037) | 0.031 | -0.0001 [-0.001, 0.001] | 0.876 | — | — | — |
| JScore_SL_2 | 607 | 329 | 0.12 (0.036) | 0.001 | 0.003 (0.042) | 0.941 | -0.08 (0.037) | 0.031 | -0.08 (0.037) | 0.032 | 0.0004 [-0.010, 0.010] | 0.941 | — | — | — |
| Light_PC1 | 486 | 249 | 0.31 (0.042) | 0.000 | -0.004 (0.038) | 0.908 | -0.075 (0.038) | 0.050 | -0.073 (0.04) | 0.064 | -0.0014 [-0.025, 0.022] | 0.908 | — | — | — |
| Light_PC2 | 486 | 249 | 0.136 (0.041) | 0.001 | -0.02 (0.038) | 0.587 | -0.075 (0.038) | 0.050 | -0.072 (0.038) | 0.061 | -0.0028 [-0.013, 0.007] | 0.592 | — | — | — |
| Light_PC3 | 486 | 249 | 0.007 (0.038) | 0.845 | -0.001 (0.039) | 0.969 | -0.075 (0.038) | 0.050 | -0.075 (0.038) | 0.050 | -0.0000 [-0.001, 0.001] | 0.970 | — | — | — |
| **Light_PC4** | **486** | **249** | **-0.4 (0.039)** | **0.000** | **0.132 (0.04)** | **0.001** | **-0.075 (0.038)** | **0.050** | **-0.022 (0.041)** | **0.587** | **-0.0529 [-0.086, -0.020]** | **0.002** | **-0.0529 [-0.080, -0.028]** | **<0.001** | **66.2** |

**Note**: Sobel test p < 0.10 used as screening threshold for bootstrap mediation testing. Mediators meeting this threshold are shown in bold.

**References**

[1]	Migueles JH, Rowlands AV, Huber F, Sabia S, van Hees VT. GGIR: A Research Community–Driven Open Source R Package for Generating Physical Activity and Sleep Outcomes From Multi-Day Raw Accelerometer Data. Journal for the Measurement of Physical Behaviour. 2019;2(3):188-96. <https://doi.org/10.1123/jmpb.2018-0063>.

[2]	Guo W, Leroux A, Shou H, Cui L, Kang SJ, Strippoli M-PF, et al. Processing of Accelerometry Data with GGIR in Motor Activity Research Consortium for Health. Journal for the Measurement of Physical Behaviour. 2023;6(1):37-44. <https://doi.org/10.1123/jmpb.2022-0018>.

[3]	Kang SJ, Leroux A, Guo W, Dey D, Strippoli M-PF, Di J, et al. Integrative Modeling of Accelerometry-Derived Sleep, Physical Activity, and Circadian Rhythm Domains With Current or Remitted Major Depression. JAMA Psychiatry. 2024;81(9):911-8. <https://doi.org/10.1001/jamapsychiatry.2024.1321>.

[4]	Hand AJ, Stone JE, Shen L, Vetter C, Cain SW, Bei B, et al. Measuring light regularity: sleep regularity is associated with regularity of light exposure in adolescents. Sleep. 2023;46(8). <https://doi.org/10.1093/sleep/zsad001>.

[5]	R Core Team. A language and environment for statistical computing Vienna, Austria: R foundation for statistical computing. In; 2022.

[6]	Bates D, Mächler M, Bolker B, Walker S. Fitting linear mixed-effects models using lme4. Journal of statistical software. 2015;67:1-48.

[7]	Shrout PE, Bolger N. Mediation in experimental and nonexperimental studies: new procedures and recommendations. Psychological methods. 2002;7(4):422.

[8]	Hayes AF. Beyond Baron and Kenny: Statistical Mediation Analysis in the New Millennium. Communication Monographs. 2009;76(4):408-20. <https://doi.org/10.1080/03637750903310360>.
